# Leveraging infectious disease models to interpret randomized controlled trials: controlling enteric pathogen transmission through water, sanitation, and hygiene interventions

**DOI:** 10.1101/2022.04.28.22274441

**Authors:** Andrew F. Brouwer, Marisa C. Eisenberg, Kevin M. Bakker, Savannah N. Boerger, Mondal H. Zahid, Matthew C. Freeman, Joseph N.S. Eisenberg

## Abstract

Randomized controlled trials (RCTs), which evaluate hypotheses in specific contexts, are often considered the gold standard of evidence for infectious disease interventions, but their results cannot immediately generalize to other contexts. Mechanistic models are one approach to generalizing findings between contexts, but infectious disease transmission models are not immediately suited for analyzing RCTs, since they often rely on time-series surveillance data that is rarely collected by RCTs. We developed a modeling framework to explain the main outcome of an infectious disease RCT—relative risk—and applied it to a water, sanitation, and hygiene (WASH) RCT. This model can generalize the RCT results to other contexts and conditions. We developed this compartmental modeling framework to account for key WASH RCT factors: i) transmission across multiple environmental pathways, ii) multiple interventions applied individually and in combination, iii) adherence to interventions or preexisting conditions, and iv) the impact of individuals not enrolled in the study. We employed a hybrid sampling-importance resampling and estimation framework to obtain posterior estimates of mechanistic parameters and their uncertainties and illustrated our model using WASH Benefits Bangladesh RCT data (n=17,187). Our model reproduced reported diarrheal prevalence in this RCT. The baseline estimate of the basic reproduction number ℛ_0_ for the control arm (1.15, 95% CI: 1.09, 1.27) corresponded to an endemic prevalence of 13% (95% CI: 9–21%) in the absence of intervention or preexisting WASH conditions. No single pathway was likely able to sustain transmission: pathway-specific ℛ_0_s for water, fomites, and all other pathways were 0.49 (95% CI: 0.07, 0.99), 0.26 (95% CI: 0.04, 0.57), and 0.40 (95% CI: 0.02, 0.88), respectively. An infectious disease modeling approach to evaluating RCTs can complement RCT analysis by providing a rigorous framework for generating data-driven hypotheses that explain trial findings, particularly unexpected null results, opening up existing data to deeper epidemiological understanding.

**Author summary:** A randomized controlled trial (RCT) testing an intervention to reduce infectious disease transmission can provide high-quality scientific evidence about the impact of that intervention in a specific context, but the results are often difficult to generalize to other policy-relevant contexts and conditions. Infectious disease transmission models can be used to explore what might happen to disease dynamics under different conditions, but the standard use of these models is to fit to longitudinal, surveillance data, which is rarely collected by RCTs. We developed a framework to fit an infectious disease model to steady-state diarrheal prevalence data in water, sanitation, and hygiene RCTs, explicitly accounting for completeness, coverage, and compliance. Although this framework is developed with water, sanitation, and hygiene interventions for enteropathogens in mind, it could be extended to other disease contexts. By leveraging existing large-scale RCT data sets, it will be possible to better understand the underlying disease epidemiology and investigate the likely outcomes of policy-relevant scenarios. Ultimately, this work can be incorporated into decision making for public health policy and programs.

## Introduction

Randomized controlled trials (RCTs) are an important source of scientific evidence in the field of epidemiology. However, while they provide high-quality evidence on the causal impact of interventions, they are also inflexible. They answer only a specific set of questions, i.e., they determine whether and to what extent the interventions led to a detectable reduction in disease burden. RCTs are not designed to decipher null results, i.e., when the expected health benefits are not realized, they provide little information on important outstanding explanatory questions. Systematic reviews and meta-analysis are often employed to increase the collective power of a collection of related trials. However, these approaches are not able to account for factors that could provide insight into the mechanism of transmission reduction, such as variability in baseline exposures (conditions), differences in background rates of infection or disease burden across study sites, heterogeneous interventions, or differences coverage and adherence of the intervention. As a result, expensive and time-consuming trials result in narrow causal inference and may provide minimal actionable data to policy makers eager to understand how findings could and should be generalized and applied to different contexts.

Infectious disease transmission models are an important tool that allow us to make inferences about disease spread and dynamics within a population. The standard use of transmission models for inference largely relies on time-series incidence data, usually in the form of passive surveillance, where the dynamic signature of time-series data can provide valuable information about an infectious disease transmission system. However, longitudinal surveillance is only one of many types of data that can provide insight into the epidemiology of infectious diseases. RCTs are concerned with comparisons between individuals across intervention arms in the form of relative risks and are generally do not include the collection of longitudinal surveillance data. If data from large-scale RCTs could be leveraged to inform mathematical models, it would open up a wealth of data that could be used to better understand epidemiological findings. Mechanistic models can play a pivotal role in developing intervention strategies and estimating what health benefits can be expected under various scenarios.

Decades of RCTs and observational studies have been conducted in the field of water, sanitation, and hygiene (WASH), constituting a wealth of data on the epidemiology of enteric disease. WASH improvements have been responsible for major public health gains by greatly reducing diarrheal disease burden caused by a variety of enteropathogens. Reducing the diarrheal burden can also result in improved nutrition and reduced stunting of child growth [1]. Transmission of the enteropathogens responsible for diarrheal disease can occur through multiple, interconnected pathways by which a susceptible person may come in contact with pathogens shed (in feces) by an infected individual. These pathways are often visualized as an “F-diagram,” illustrating some of the potential transmission routes (e.g., feces, fingers, fomites, fluids, and food) and potential points of intervention [2] (Fig. 1). Recent large scale, well-powered intervention trials have not shown the expected health benefits of WASH interventions, namely reduced diarrhea and improved growth [3–9]. Moreover, while WASH improvements such as latrines have demonstrable efficacy at the household level, they have not yielded the expected health improvements at the community level [10]. This failure of community-level efficacy is likely due to a combination of three factors: the interventions did not sufficiently reduce transmission across the multiple pathways (completeness), the coverage did reach a sufficient level to induce herd protection (coverage), or the adherence to the interventions was not sufficiently great (compliance) [11].

**Fig 1.**
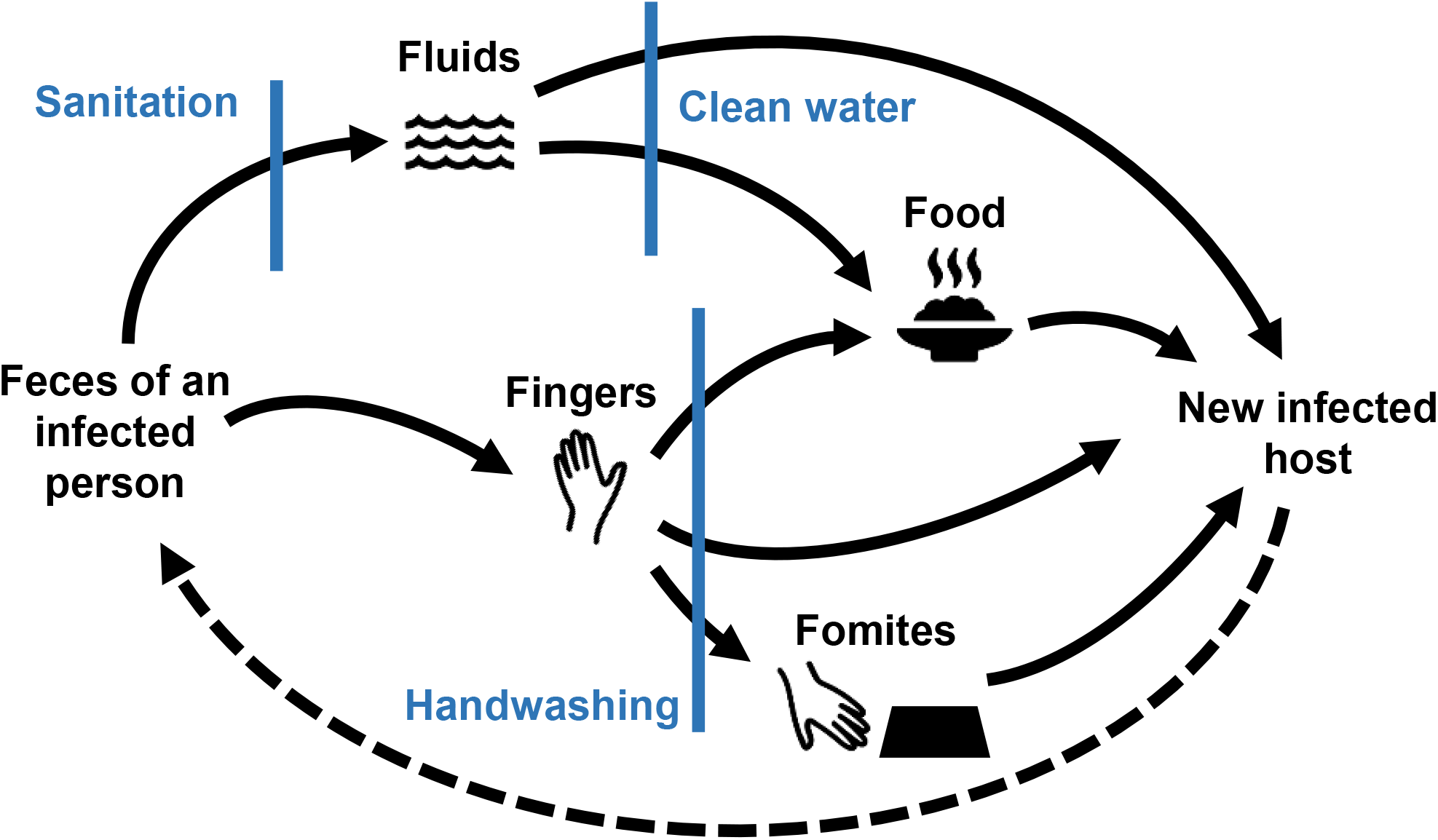
F-diagram. Simplified “F-diagram” illustrating different transmission pathways [2]. The blue bars show how specific types of interventions may interrupt transmission along these pathways. The dotted line (not traditionally included in the F-diagram), highlights that infected individuals continue to contribute to environmental contamination.

Here, we present a transmission modeling framework that develops a mechanistic understanding of the WASH RCT data, explicitly accounting for completeness, coverage, and compliance. To this end, we developed a compartmental infectious disease modeling framework and parameter estimation approach to analyzing RCT data. This analytical framework was designed to incorporate the data underlying relative risk estimates and other contextual data collected by an RCT to calibrate a mechanistic model. A calibrated model could then be used to examine counterfactual questions (i.e., “what would have happened if … ?”) that provide the basis to generate hypotheses on mechanisms behind the observed relative risk estimates. We applied our framework and approach using data from WASH Benefits Bangladesh [6, 12], a large, seven arm, cluster-randomized controlled trial on the impact of WASH and nutrition on diarrhea and child growth outcomes. Specifically, the model generated prevalence estimates from steady-state simulations accounting for multiple environmental pathways, individual adherence to multiple WASH interventions, preexisting WASH conditions, and contribution of the subpopulation that was not enrolled into the RCT study to transmission. We used sampling-importance resampling to quantify the uncertainty in key parameters of interest, such as intervention efficacy and the relative contribution of different environmental pathways.

## Methods

### Randomized controlled trial design

While no two *randomized controlled trials* (RCTs) are exactly alike, they often have similar characteristics. The objective of an infectious-disease-related RCT is to determine the effectiveness of an intervention, and, if the interventions are effective, the RCT may estimate pathway-specific attributable risks. RCTs are the one true experimental tool in epidemiology and are often considered the gold standard of scientific evidence [13–15] because the intervention is randomly assigned, thereby addressing confounding in the design phase of the study. Participants are randomly assigned to one or more groups, or *arms*. An *intervention* is applied to one arm (e.g., a water treatment device and health promotion visits from study staff), while the comparison group, the *control* arm, receives either no intervention, a placebo, or an alternative intervention (e.g., standard care, such as health promotion visits but no water treatment device). Typically, specified measurements are taken at a baseline time point prior to intervention and at one or more follow-up time points after the interventions are applied. More complicated intervention designs are possible, including multiple interventions that are compared to the control or each other. In the case of WASH RCTs, different interventions are used to target different environmental pathways (Fig. 1). Multiple interventions applied together are used to evaluate the *completeness* of the interventions in blocking transmission.

RCT data are often analyzed using one of two strategies. An *intention-to-treat* (ITT) approach analyzes data by grouping individuals by their intervention assignment. The ITT approach results in a measure of intervention *effectiveness*, reflecting real-world usage. Effectiveness can differ from the true underlying potential *efficacy* of the intervention used under ideal conditions because of imperfect intervention *fidelity*, i.e., delivery of the intervention, or imperfect intervention *compliance/adherence*, i.e., the use of the intervention by the recipient as intended. A *per protocol* approach to analysis of the RCT removes participants who did not receive or did not adhere to their intervention assignment. This approach can provide an estimate of intervention effectiveness that is closer to the true efficacy, but the benefits of randomization are lost. The magnitude of risk reduction associated with an intervention depends not only on the its efficacy at blocking transmission, but also on the endemic pathogen prevalence, the susceptibility of the target population, the quality of the preexisting WASH *conditions* (e.g., an improved latrine may have greater benefits if it replaces open defecation compared to if it replaces an unimproved latrine), and other factors.

Interventions in a WASH RCT are usually delivered in a way that would be practical for subsequent programs to implement at a greater scale, which means that interventions are usually provided at the household, village, school, or district level, rather than the individual level. Moreover, rather than randomizing individuals to treatment arms, RCTs may employ a cluster-randomized design to randomize larger groups, or *clusters*, because WASH interventions likely affect those beyond the intended target, i.e., they have both direct effects on the participant and indirect effects on the community through reduced infection pressure. The fraction of the population in an area that receives the interventions is *coverage* of the trial; in some trials, all individuals in a cluster are receive the intervention, and in other trials, only a subset or only eligible individuals receive the intervention.

### WASH randomized clinical trial data

For the purposes of developing an analysis framework, we lay out our assumptions about what is included in a WASH RCT data set reporting on diarrheal outcomes.

- Random allocation occurs at the cluster level, and we know which intervention arm and cluster each individual is in.
- We know whether each enrolled individual has preexisting WASH conditions substantively equivalent to the intervention. If not, we assume that the preexisting WASH conditions are not equivalent to the intervention.
- The preexisting WASH conditions of people not enrolled the study are approximated by the conditions of people in the control arm at that point in time.
- We know whether each individual received and adhered to their intervention (resulting in a per-protocol-like approach). If not, we assume that all individuals received and adhered to their intervention (resulting in an ITT-like approach).
- We know the fraction of the population enrolled in the study (e.g., through a census). If not, we will estimate it.
- The measured outcome for each enrolled individual is a self-report (or guardian report) of whether they recently had (all-cause) diarrhea (e.g., in the past seven days).

### Mathematical modeling framework

Our framework is comprised of a transmission model representing WASH interventions applied to a subset of a population and a parameter estimation approach designed to fit data to steady-state modeling simulations. Our model structure incorporates several important assumptions. First, the model reflects a cluster RCT design where each cluster includes both the fraction of the population enrolled in the study and the remainder that is not. Second, the model explicitly accounts for adherence and fidelity (i.e., the extent to which the intervention is implemented as intended), and therefore it uses a per-protocol-like approach where only those who received and are adhering to the intervention are considered in the protected group. Third, the model does not distinguish between those adhering to the intervention and those that have preexisting WASH conditions that block the same transmission pathway as the intervention. For example, we do not distinguish between an preexisting improved latrine and one provided by the RCT. Finally, everyone within a given cluster is modeled as sharing the same environment. These assumptions are specific to our WASH RCT analysis and the WASH Benefits study design. Other study designs and infectious disease systems would necessitate an alternative model structure. Below, we describe the model structure and the parameter estimation approach.

#### Developing the single-intervention SISE–RCT model

The basis of our modeling framework is the environmental infectious disease transmission model, the SIRE (susceptible, infectious, recovered, environment) model [16, 17]. This model tracks the fraction of individuals that are susceptible (*S*), infectious (*I*), or recovered (*R*), as well as the concentration of pathogens in the environment (*E*). Pathogen transmission occurs when susceptible people contact the environment. Here, the contact rate, contact volume, and pathogen infectiousness are all combined into a single transmission parameter *β* [18]. Infectious individuals recover at rate *γ* and shed pathogen into the environment at rate *α*. Pathogens decay in the environment at rate *ξ*. The differential equations governing this model are

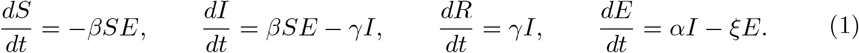

The basic reproduction number of the SIRE model, which is a measure of the epidemic potential of the system, is 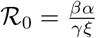

For the RCT framework, we modified the above model in several ways. First, because the WASH RCT outcome measure is all-cause and not pathogen-specific diarrhea, we used an SIS (susceptible–infectious–susceptible) framework. Individuals return to the susceptible compartment after their infectious period rather than progressing to a recovered compartment, as they remain susceptible to other enteropathogens. An SIS model can have an endemic equilibrium, where the disease remains prevalent at some level in the population. For many SIS models, the value of this endemic equilibrium is 1 − 1/ℛ_0_, so that the larger the ℛ_0_, the more prevalent the disease at equilibrium.

Second, we explicitly modeled two populations that share a single environment, i.e., a single cluster (Fig. 2). The first population, denoted by the subscript +, received and is adhering to an intervention or has a substantively equivalent preexisting WASH condition (e.g., already has an improved latrine in a trial providing improved latrines).

**Fig 2.**
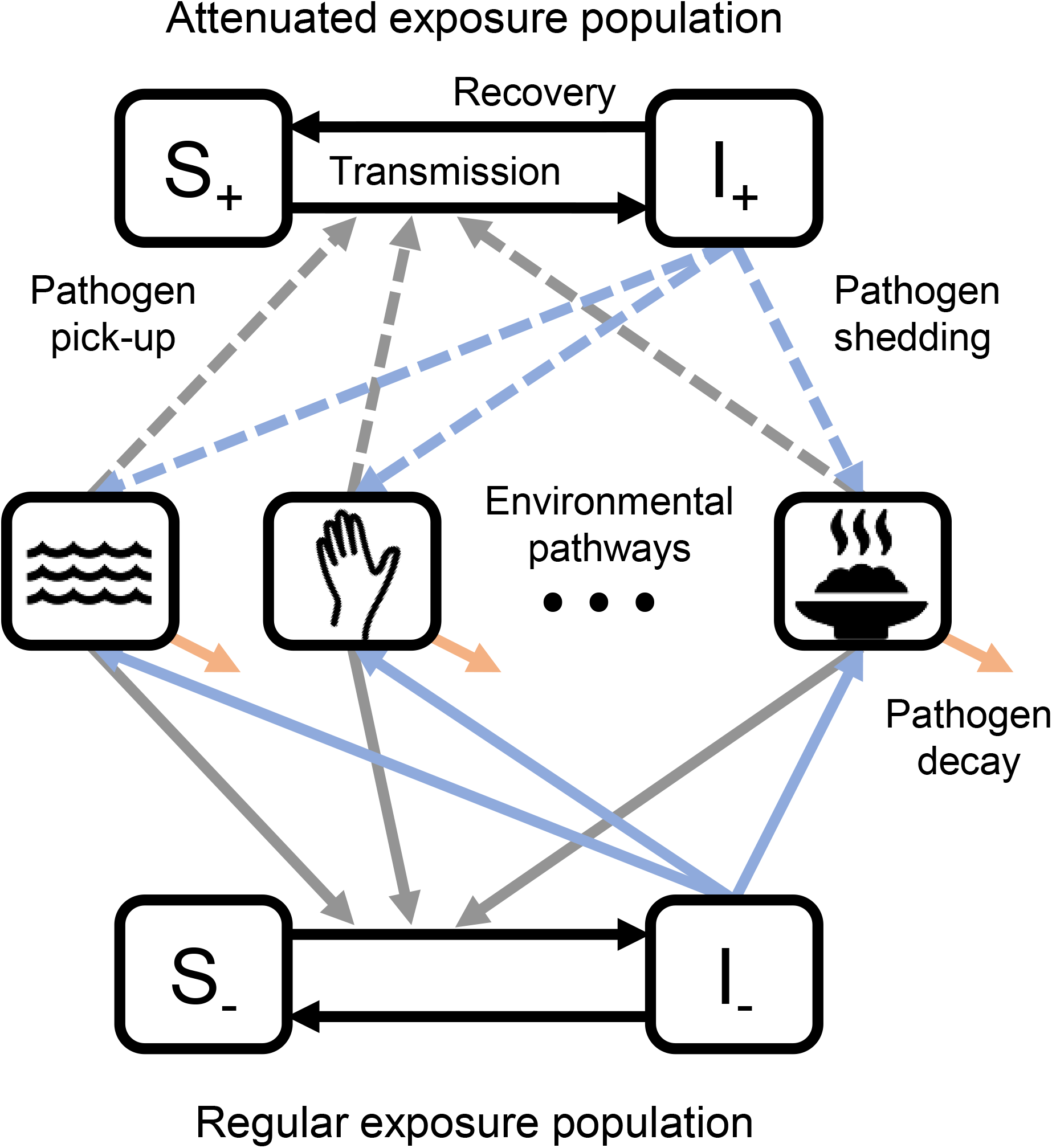
Environmental transmission in an intervention study. The shared environment has multiple environmental pathways that correspond to transmission routes depicted on a traditional F-diagram. The first population (subscript +) adheres to an intervention or has preexisting WASH conditions that provide the same protection as the intervention; this population has attenuated exposure to and attenuated shedding into the environment (dashed lines). The second population (subscript -) is not covered by the study or does not adhere to the intervention and has no preexisting WASH conditions; this population has regular exposure and shedding (solid lines).

For brevity going forward, “preexisting WASH condition” will refer to a preexisting WASH condition that is substantively equivalent to the corresponding intervention. Accordingly, this population has attenuated exposure to or shedding into the environment. The second population, denoted by the subscript −, is not enrolled in the study, or has not received or is not adhering to the intervention and has no preexisting WASH condition. This population has regular exposure and shedding, although it may receive indirect benefits of the intervention through reduced environmental contamination. We use this framework for clusters both in the intervention arm and in the control arm. In the intervention arm, all study households receive the intervention, though not all adhere to the intervention, while local, non-study households do not receive the intervention but may have preexisting WASH conditions that are equivalent to the intervention. In the control arm, no one receives the intervention, and both study households and non-study households may or may not have preexisting WASH conditions.

Let *ρ* be the fraction of those in a cluster (i.e., with a shared environment) that are adhering to the intervention or an equivalent preexisting WASH condition. Let *ρ*_0_ be the fraction of those *not* in the study that have a preexisting WASH condition equivalent to the intervention. Let *ω* be the fraction of the population in the study (coverage). The size of the population adhering to intervention or with a preexisting condition is *ωρ* + (1 *− ω*)*ρ*_0_, and the size of the population not adhering to the intervention or preexisting condition population is *ω*(1 − *ρ*) + (1 − *ω*)(1 − *ρ*_0_).

Third, we extend the model to account for multiple modes of environmental transmission, *E*_1_, *E*_2_, …, *E*_*n*_. The modes represent different pathways on the F-diagram (Fig. 1), such as fluids and fomites. We always include an “other” pathway to account for transmission pathways not intervened on. We define which part of the transmission process is impacted by each of the modeled interventions. An intervention may reduce the transmission rate by reducing the number of pathogens contacted or the susceptibility of the individual. We denote the relative transmission by 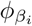, where subscript *β*_*i*_ denotes an impact on transmission from environment *i*, and thus the intervention efficacy by 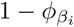. For example, if an intervention has 80% efficacy in reducing transmission, the magnitude of the remaining transmission is 20% of the original. The intervention may instead reduce the shedding of pathogen to the shared environment, with efficacy 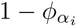, where *α*_*i*_ denotes the impact on shedding into environment *i*.

This expanded model (SISE–RCT) has 4 equations representing the human population and *n* equations representing the environmental pathways (Fig. 2). These equations are provided in the supporting information (Eq. S1).

#### Identifiability and reparameterization

The initial goal of this work is to determine what model parameter values are consistent with the observed RCT data. As we discuss in a later section in more detail, we connect the model to the data through the modeled steady-state diarrheal prevalence. To estimate the value of a parameter from the data, it must be identifiable from steady-state prevalence, that is, the parameter must have a unique value associated with a given steady state.

To solve for the steady-state values of the model, we first set each *dE*_*i*_*/dt* equation in the SISE–RCT model (Eq. (S1)) to 0 (a quasi-steady-state assumption), solve for each *E*_*i*_, and substitute those expressions into the remaining equations (Eq. (S3)). When these resulting equations are at steady state, for each pathway *i* the model parameters *β*_*i*_, *α*_*i*_, *ξ*_*i*_, and *γ*_*i*_ only appear together in a certain parameter combination and thus are not separately identifiable from steady-state data. We define these identifiable combinations of those four parameters as the pathway-specific reproduction numbers,

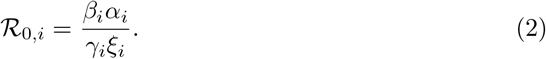

The magnitude of each ℛ_0,*i*_ relates to the strength of transmission along pathway *i*. Note, too, that ℛ_0_ = ∑_*i*_ ℛ_0,*i*_ is the basic reproduction number of the system in Eq. (S1) in the absence of any efficacious intervention, i.e., each 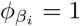 and 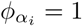. For parameter estimation, it is convenient to express the strength of the transmission pathways as relative to the total ℛ_0_, that is ℛ_0,*i*_*/ℛ*_0_.

The equations Eq. (S3) resulting from the quasi-steady-state assumption on the environmental states and reparameterization (in terms of *βα/ξ* and *γ*) still include dynamics for the susceptible and infectious states, and there is not a closed-form solution for the steady state values. However, we can use numerical simulation to calculate the steady state solutions as a function of the ℛ_0,*i*_ only and not their constituent parameters using the following observation. If we divide the right hand side of each equation in Eq. (S3) by *γ*, the equations are only parameterized by the ℛ_0,*i*_, and this set of equations has same steady state values as the original equations (Eq. (S1)), which is our goal. (Note that the transient dynamics of these equations are no longer biologically meaningful).

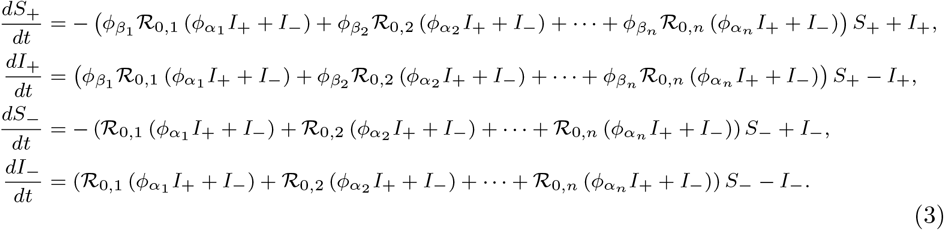

#### Adjusting for arm- and time-point-specific variation in force of infection

If an RCT takes multiple measurements (even just a pre-intervention and post-intervention measurement), it may be necessary to adjust the model for systematic differences in the force of infection across those time points. For example, external stressors, such as weather, may be vary across time points, as may important demographic factors such as the age distribution.

While succesful randomization should render the baseline disease prevalence across arms to be essentially the same, therefore negating the need to adjust for covariates when estimating relative risks, when fitting the data in mechanistic models it may be beneficial to account for actual differences across arms in potential confounders or external factors to improve our model fits to the data.

We account for any systematic differences in ℛ_0_ across time points *t* = 1, …, *T* and between the control (*a* = 1) and intervention arms (*a* = 2), by defining time- and arm-specific relative reproduction number parameters *η*_*t*_ and *η*_*a*_, relative to the basic reproduction number baseline and the control arm, respectively. Thus, we can represent the diarrheal disease pressure at time *t* in arm *a* as 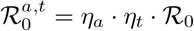, where *η*_*t*=1_ = 1 at baseline and *η*_*a*=1_ = 1 for the control arm.

#### Summary of model parameters

We have now defined all parameters needed to specify the SISE-RCT model, which will be used to calculate cluster- and intervention vs non-intervention population-specific steady-state prevalence values. These parameters consist of i) the overall basic reproduction number ℛ_0_; ii) pathway-specific basic reproduction numbers relative to the total basic reproduction ℛ_0,*i*_*/ ℛ*_0_, one for each environmental pathway *i*; iii) efficacy parameters indicating the effect of the intervention on transmission *ϕ*_*β,i*_ and shedding *ϕ*_*α,i*_ for each pathway *i*; iv) the arm- and time-specific relative basic reproduction numbers *η*_*t*_ and *η*_*a*_; and v) the coverage *ω* (if unknown). Collectively, we denote this set of model parameters {ℛ_0_, {ℛ_0,*i*_*/ℛ*_0_}, {*ϕ*_*β,i*_}, {*ϕ*_*α,i*_}, {*η*_*a*_}, {*η*_*t*_}, *ω*} as ***θ***, where internal brackets denote a set of multiple parameters.

#### Expanding the model framework to an RCT with multiple intervention arms

This single intervention framework can be expanded to account for multiple interventions within a single RCT. In the context of WASH, an RCT may have water, sanitation, and hygiene interventions arms, as well as arms evaluating some combination of these interventions. The RCT may also include interventions indirectly associated with WASH, such as nutrition. For an RCT with multiple interventions, the number of modeled populations is 2 raised to the power of distinct interventions. For example, to model an RCT testing 3 interventions in some number of combinations, we model each cluster as partitioned into 8 populations denoted by whether individuals are independently adhering to each of the three interventions (or preexisting conditions): *I*_000_, *I*_100_, *I*_010_, *I*_001_, *I*_110_, *I*_101_, *I*_011_, *I*_111_, where a 1 in the subscript represent that individuals in the group receive and adhere to the respective intervention or preexisting WASH condition and a 0 represents that they do not. We refer to these populations as *adherence groups*. Note that the number of adherence groups does not depend on the number of arms in the RCT. The RCT may or may not be investigating any given combination of interventions, but because individuals may or may not be adhering to each intervention or the associated preexisting condition, all adherence groups are modeled in all clusters in all arms. We replace the single intervention adherence fraction *ρ* by a vector denoting the fraction of the population in each of adherence groups ***ρ***. We denote the specific distribution of adherence groups in cluster *j* as ***ρ***_*j*_. In a cluster, the population not enrolled in the study also has a distribution of adherence groups, denoted ***ρ***_**0**_, which only includes adherence to preexisting conditions. Because we do not have a measure of ***ρ***_**0**_, as detailed earlier, we assume that it follows the mean baseline distribution of preexisting conditions and is the same in all clusters.

The differential equation model defines a set of steady state prevalence values ***π***_*j*_ among the adherence groups in cluster *j* as a function of the model parameters ***θ*** and the distribution of adherence groups among people in the study ***ρ***_*j*_ and not in the study ***ρ***_0_. We denote the steady state prevalence values in this cluster ***π***_*j*_(***θ, ρ***_*j*_, ***ρ***_0_) as a function of the model parameters. We are interested in prevalence estimates for each observation *k*, i.e., for a given individual at a given time point. Because ***ρ***_*j*_ is known from the data for any observation *k* in cluster *j* and as ***ρ***_0_ is assumed to be equal to that of the control group, we can drop explicit dependence on these quantities when we denote the modeled prevalence for observation *k* as a function of the model parameters, *π*_*k*_(***θ***). Note that all individuals in the same adherence group at a given time will have the same associated modeled prevalence.

### Parameter estimation

#### Statistical likelihood

We connect the model parameters to the data through a goodness-of-fit function, the likelihood *ℒ*. Because self-reported diarrhea is a binary outcome, we use a Bernoulli likelihood. As defined above, let ***θ*** be the vector of model parameters, *π*_*k*_ be the modeled prevalence corresponding to observation *k*, and *x*_*k*_ be the indicator of diarrhea for observation *k*. Then, the likelihood is given as

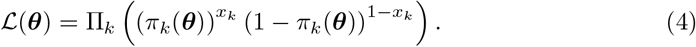

#### Sampling-importance resampling

We expect that in most RCT contexts, most of the model parameters ***θ*** will not be strongly determined by the available data, i.e., will have large uncertainty around their values. For example, a 30% water intervention efficacy may explain the data as well as a 70% efficacy, for certain values of the other parameters. In this situation, the likelihood space may be flat or multi-modal. Accordingly, it may not be possible to determine maximum-likelihood estimates, and asymptotic confidence intervals may not be representative of the true uncertainty.

Instead, we take a sampling approach to understand the distribution and uncertainty of the parameter values that explain the RCT data. First, we sample a large number of parameter sets from a prior distribution (such as a uniform distribution, as we use here). Then, we evaluate the importance of these samples by calculating their likelihood (Eq. 4). Finally, we resample the parameter sets, weighting the parameter sets by their likelihoods, to generate posterior distributions of our parameters. This Bayesian approach is known as sampling-importance resampling [19, 20].

#### Hybrid sampling–estimation approach

Instead of sampling all parameters in the sampling step of the sampling-importance resampling procedure, we sample a subset of the parameters, treat them as fixed, and estimate the remaining parameters. This approach creates a hybrid sampled–estimated set of initial parameters that we subsequently calculate the likelihoods for and resample from. The advantage of this approach is that it reduces the number of parameter samples needed by preventing a parameter set that could otherwise fit the data well from being discarded because an identifiable parameter was sampled poorly. This hybrid approach requires that the parameters to be estimated be practically identifiable, given fixed values of the sampled subset of parameters. Because ℛ_0_ is closely tied to steady state prevalence, there will be a best-fit value of ℛ_0_ for a given set of coverage, efficacy, and pathway-specific relative ℛ_0_ parameters. Thus, for each sample of the coverage, efficacy, and relative pathway parameters, we find the maximum-likelihood value of ℛ_0_ and arm- and time-specific relative basic reproduction numbers *η*_*a*_ and *η*_*t*_. This maximum likelihood is then associated with the sampled parameter set. Once a likelihood is established for each sampled–estimated parameter set, the posterior distribution can be estimated using sampling-importance resampling. More formally, we take the following steps.

1. Define sets of parameters to be sampled 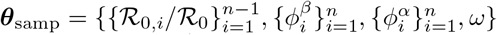 and parameters to be estimated 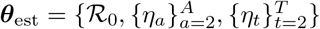, taking advantage of any degeneracies (e.g., we do not need to estimate 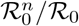 if we know ℛ_0_ and 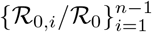.
2. Define parameter sample sets of values of 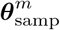 for *m ∈ M* using a multivariate uniform distribution or a more efficient algorithm such as Latin hypercube sampling [21] or a Sobol sequence [22].
3. For each 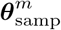, use an optimization algorithm to minimize the negative log-likelihood as a function of ***θ***_est_, and set 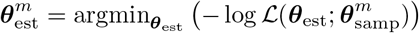
4. Define unnormalized weights *ν*_*m*_ for each 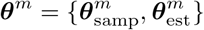 as the corresponding likelihood value divided by the probability of the sample in the originating (uniform) distribution *ν*_*m*_ = *ℒ*(***θ***^*m*^)*/*(1*/M*). Define normalized weights 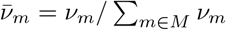. It may be preferable to directly compute the normalized weights as

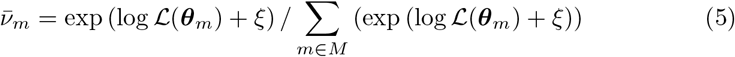

where *ξ* = min_*m∈M*_ (−logℒ(***θ***_*m*_)) is the minimum negative log-likelihood among the parameter samples.
5. Sample *N* parameter sets, with replacement, from {***θ***^*m*^} using the normalized weights 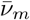.
6. These *N* parameter sets approximate the posterior distribution of ***θ*** and thus describe our knowledge of about the uncertain parameter. Summarize these distributions using histograms.

We summarize the hybrid sampling–estimation approach in Figure 3.

**Fig 3.**
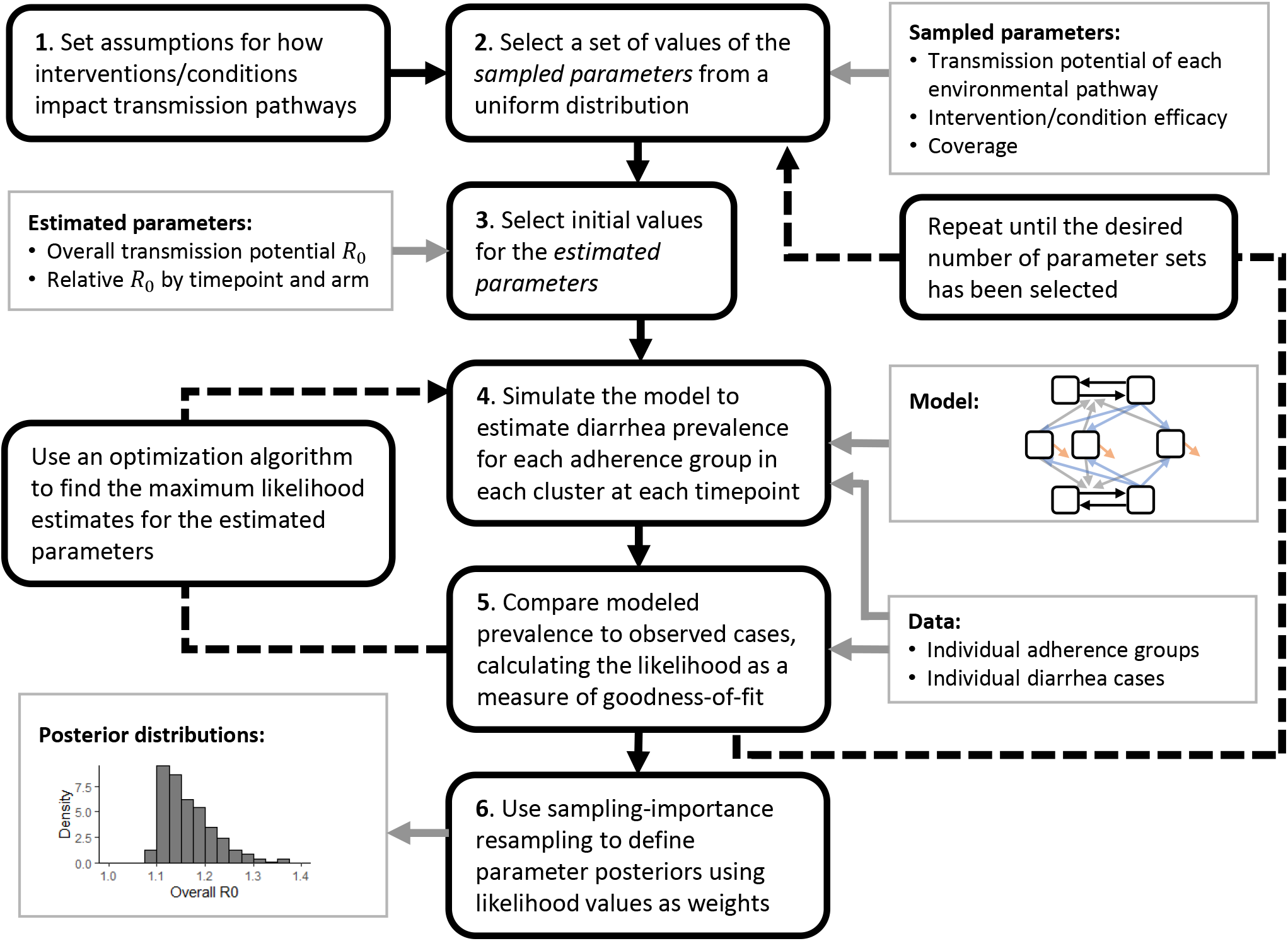
Schematic. Schematic of the hybrid sampling–estimation approach to estimating model parameters from Water, Sanitation, and Hygiene (WASH) randomized controlled trials (RCT) data.

### WASH Benefits Bangladesh

We demonstrate the modeling framework using data from the WASH Benefits Bangladesh RCT [6, 12]. WASH Benefits Bangladesh measured diarrheal prevalence in children (as well as multiple child growth measures, although we do not consider those outcomes here) at each of three time points (baseline, midline, endline). Households in the study area are typically organized into compounds in which a patrilineal family shares a common space and resources, such as a water source and latrine. A total of 5551 compounds were enrolled, contingent on having a pregnant woman in their second trimester during the enrollment period. The study followed one or more target children born after baseline, as well as any siblings who were under age 3 at baseline. These compounds were grouped into 720 clusters. Each cluster was assigned to one of 7 arms testing combinations of 4 interventions: water chlorination (W), a double-pit, pour flush improved latrine (S), handwashing with soap and water (H), and supplementary nutrition sachets (N). Of the 720 clusters, 180 were assigned to the control arm (C), while 90 were assigned to each of the water (W), sanitation (S), handwashing (H), nutrition (N), combined water, sanitation, and handwashing (WSH), and all interventions (WSH-N) arms. Specific details on trial design, intervention specifics, and results may be found elsewhere [6, 12]. Because our analysis was a secondary analysis of deidentified data, it is not regulated as human subjects research.

For the purposes of assigning individuals to intervention adherence groups for the model, we classified individuals at each time point in each arm as using or not using each intervention or preexisting WASH condition using the following indicators.

- W: Free chlorine was detected in stored water.
- S: Latrine was present and had a functional water seal.
- H: Primary handwashing location was present with available water and soap.
- N: At least 50% of expected nutrition sachets were reported as being consumed.

Each of these indicators denote WASH or nutrition conditions that impact susceptibility, exposure, or shedding, as we describe in more detail below. These four indicators collectively describe 2^4^=16 adherence groups. In WASH Benefits Bangladesh, chlorine was only measured in arms with the water intervention, and we assumed that there was no use of chlorination in arms not receiving the intervention. Children not in nutrition intervention arms and non-target children in nutrition intervention arms were assumed to have not received supplemental nutrition. We removed individuals with negative reported ages (n=2), missing reported diarrhea (n=2,745), or missing in any of the four use indicators (n=2,660), which left 17,187 individual observations (76% of the original sample) over the three time points. For the remaining data, we plot the arm-specific prevalence of each of the 4 indicators (Figure 4a–d). Among target children in arms receiving the nutrition intervention, 93% reported consumption of at least 50%, the vast majority of whom (83% of target children) reported 100% consumption.

**Fig 4.**
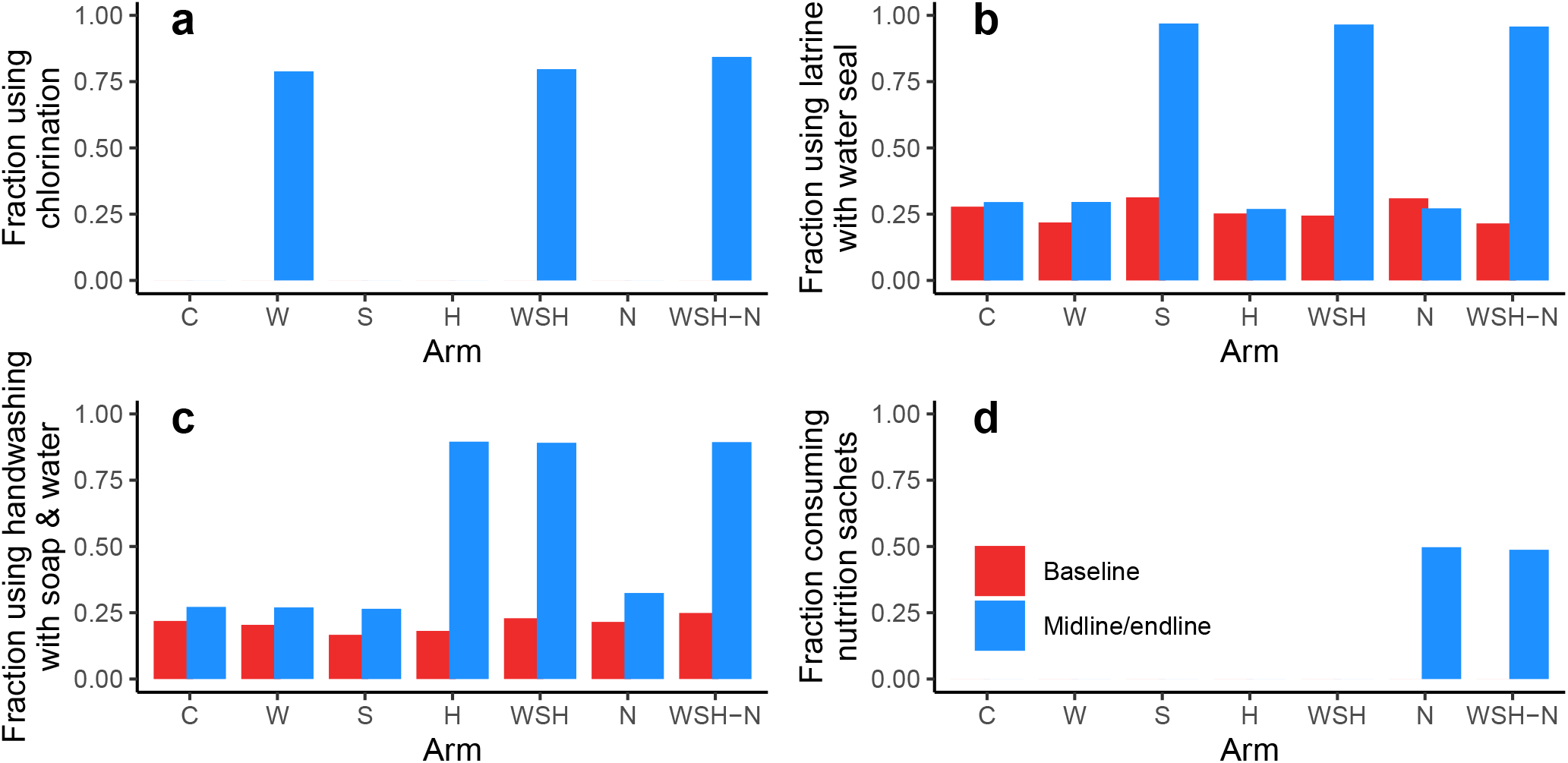
Prevalence of intervention and preexisting WASH conditions. Prevalence of (a) free chlorine, (b) latrine with water seal, (c) handwashing station with soap and water, and (d) reported 50% nutrition sachet consumption (only provided to target children). The arms are denoted by combinations of interventions, C: control, W: water, S: sanitation, H: hygiene, N: nutrition.

We connect the data to the model by making assumptions about which transmission pathways the interventions/conditions effect. We model three transmission pathways, namely water *w*, fomites *f*, and other *o* (including person-to-person). We assume chlorination (W) reduces exposure from the water pathway 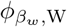, a latrine water seal (S) reduces shedding into the water pathway 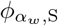, handwashing with soap and water (H) reduces exposure from the fomite pathway 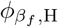, and supplemental nutrition (N) reduces susceptibility from all pathways 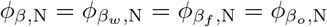 (where the first subscript, e.g., *β*_*w*_, represents the pathway-specific parameter attenuated by the intervention, and the second subscript, e.g., W, represents the intervention.) We assume transmission and shedding in all other arms and pathways are not attenuated, i.e., *ϕ*_*β*_ and *ϕ*_*α*_ are 1. In total, we considered sixteen model parameters, namely the overall ℛ_0_, pathway-specific relative ℛ_0_ parameters, 4 efficacy parameters, a coverage parameter, 6 arm-specific relative ℛ_0_ parameters, and 2 time-point-specific ℛ_0_ parameters: 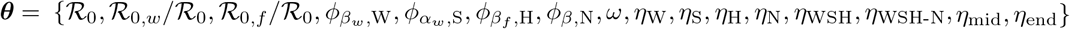. Full model equations are given in the Appendix (Eq. (S4)). We used the hybrid sampling–estimation approach (Fig. 3) to estimate posterior distributions of each of the 16 parameters. We began with prior set of *M* = 25,000 parameter sets determined by a Sobol sequence to uniformly cover the parameter space, and we present posterior distributions taking *N* = 25,000 samples (with replacement) from the prior distribution weighted by the importance (likelihood) of the prior samples.

## Results

The model of the WASH Benefits Bangladesh RCT trial reproduced the observed prevalence across the arms and time points (Figure 5a; the analogous figure with midline and endline separately plotted is given in Figure S2). The observed prevalence values are analogous and comparable to the results shown previously in Luby et al. [6], although the prevalence estimates in Figure 5a are for the subset of the population with full intervention adherence data. (See Figure S1 for a comparison of the distribution of model fits in the prior and posterior samples is provided in the supporting information). As previously reported [6], midline/endline prevalence in the W arm was comparable to the control arm, and the prevalence in the remaining arms were lower than control and similar to each other. There were no statistically significant differences in the observed baseline prevalence across the arms.

**Fig 5.**
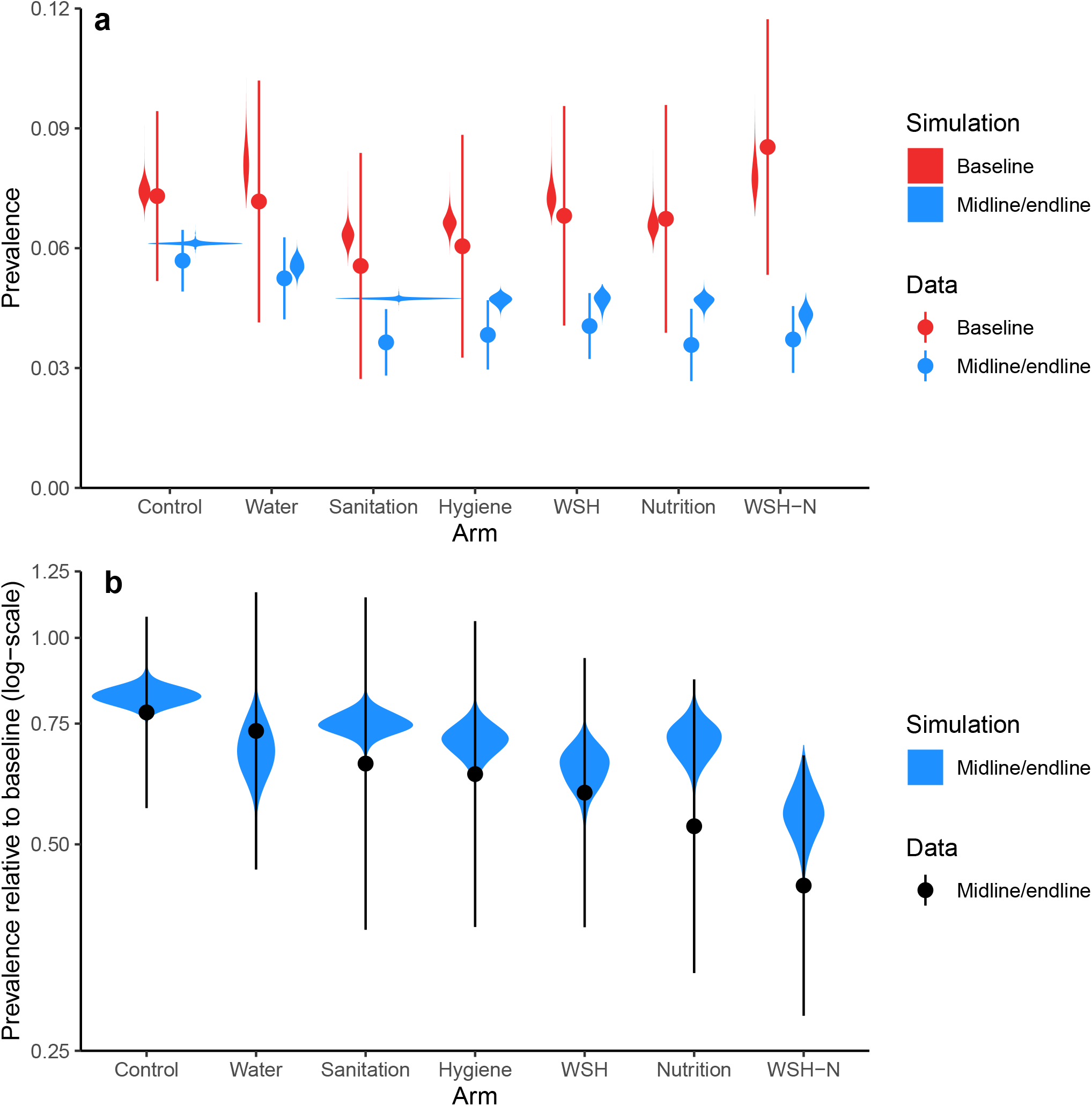
Prevalence and prevalence ratio. a) Prevalence of self-reported diarrhea (7-day recall) in WASH Benefits comparing the baseline (red) to the combined midline/endline (blue) surveys (comparable to result given in [6]), as well as posterior distributions of simulated prevalence (violine plots). b) Prevalence ratios (data and simulated) for each arm for midline/endline relative to base. In both figures, the violin plots indicate the distribution of values in the parameter posterior sample for each arm, and their areas are scaled to the number of observations.

When comparing the relative risk of midline/endline to baseline across arms, arms with combined interventions had lower point estimate prevalence ratios than arms with consituent interventions, e.g., the point estimate for the WSH-N prevalence ration was below than each of the W, S, H, WSH, and N arms. Moreover, the midline/endline to baseline prevalence ratios for all arms, including the control, were below 1. Together, these results provide a rationale for the inclusion of both arm- and time-point-specific estimates of the variation in ℛ_0_, allowing us to maximize the amount of information about the pathway and efficacy parameters.

While most of the simulation estimates were able to reproduce the data, or at least produce confidence intervals overlapping those of the data, the model estimate for the S arm does not capture the observed diarrheal prevalence in that arm at midline/endline. This discrepancy may suggest that our assumption that the efficacies of the sanitation and hygiene interventions were the same as the comparable preexisting WASH conditions is not valid for those interventions. That is, there may be a poor fit to midline/endline prevalence in those arms because, to better match the prevalence in the arms, the model would have had to also decrease the infection prevalence for individuals in the other arms that had preexisting conditions, which would have resulted in poorer fits for those arms. Alternatively, the discrepancy may be due to noise in the diarrhea recall variable [23]. The simulated prevalence for the N arm also has limited overlap with the data. This discrepancy is due, in part, to the fact the the model is fitting to individual outcomes while the plots are showing aggregrate outcomes. While diarrheal prevalence point estimate in the N arm among those who had the intervention (3.0%) was lower than that among those that did not (4.2%), this pattern did not hold in the WSH-N arm, where prevalence among those who received the intervention (3.9%) was comparable to that among those who did not (3.6%). The model cannot fit the outcomes for one group better without fitting the outcomes from another group more poorly.

The basic reproduction number ℛ_0_ corresponding to the control arm at baseline was estimated to be 1.15 (95% CI: 1.09, 1.27) (Figure 6a), which corresponds to an endemic prevalence of 13% (95% CI: 9–21%) in the absence of any preexisting WASH conditions. Estimates of timepoint- and arm-specific relative ℛ_0_ are given in the supporting information (Figure S3). The posterior distributions of the pathway-specific reproduction numbers were wide, indicating uncertainty in the estimates. The mean ℛ_0_ value for the water pathway accounted for 42% of transmission (ℛ_0,*w*_ = 0.49, 95% CI: 0.07, 0.99), while the fomite and other pathways accounted for 22% (ℛ_0,*f*_ = 0.26, 95% CI: 0.04, 0.57) and 35% (ℛ_0,*o*_ = 0.40, 95% CI: 0.02, 0.88), respectively. However, there is substantial uncertainty around the specific values. Nevertheless, the results indicate that no single pathway would be able to sustain diarrhea transmission alone.

**Fig 6.**
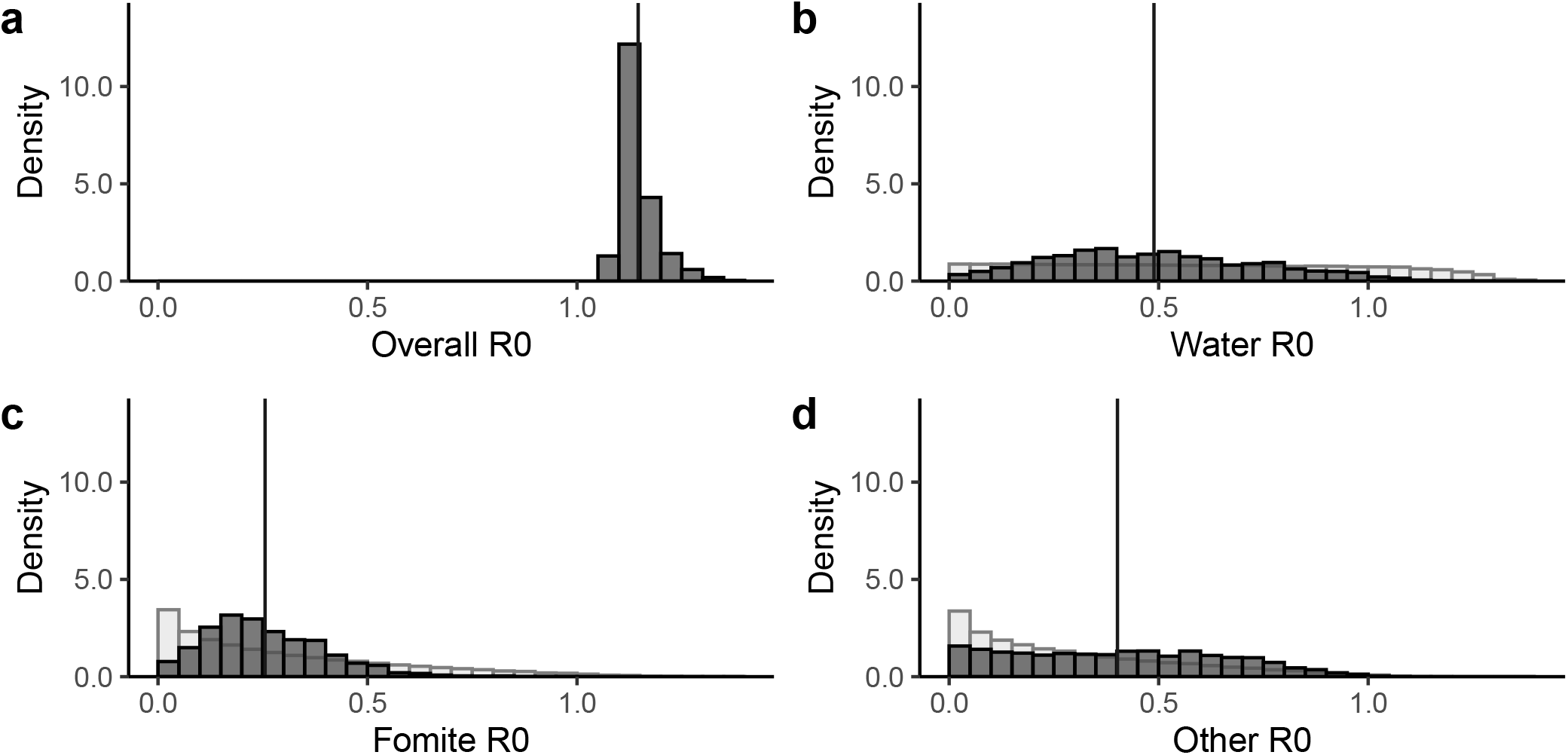
Reproduction numbers. Posterior distributions (dark grey) of the (a) overall basic reproduction number ℛ_0_, (b) the water pathway basic reproduction number, (c) the fomite pathway reproduction number, and (d) the other pathway reproduction number. Prior distributions (light grey) are given for the three sampled parameters; prior distributions are not uniform because they are products of underlying uniformly distributed parameters.

The observed prevalence values are consistent with a wide range of values for the efficacy of a) water chlorination 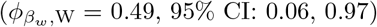, b) having a latrine with a water seal 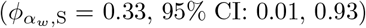, and c) handwashing with soap and water 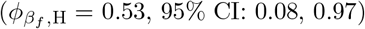, but are consistent with a comparably narrower range of efficacy of d) consumption of nutrition sachets (*ϕ*_*β*,N_ = 0.14, 95% CI: 0.01, 0.35) (Figure 7). The posterior distribution of estimated study coverage *ω* suggests that 3.8% (95% CI: 0.002, 9.3%) of the population was enrolled in the study (Figure S4); this result is an underestimate because the initial sampling range (up to 10%) did not capture the full tail of the distribution.

**Fig 7.**
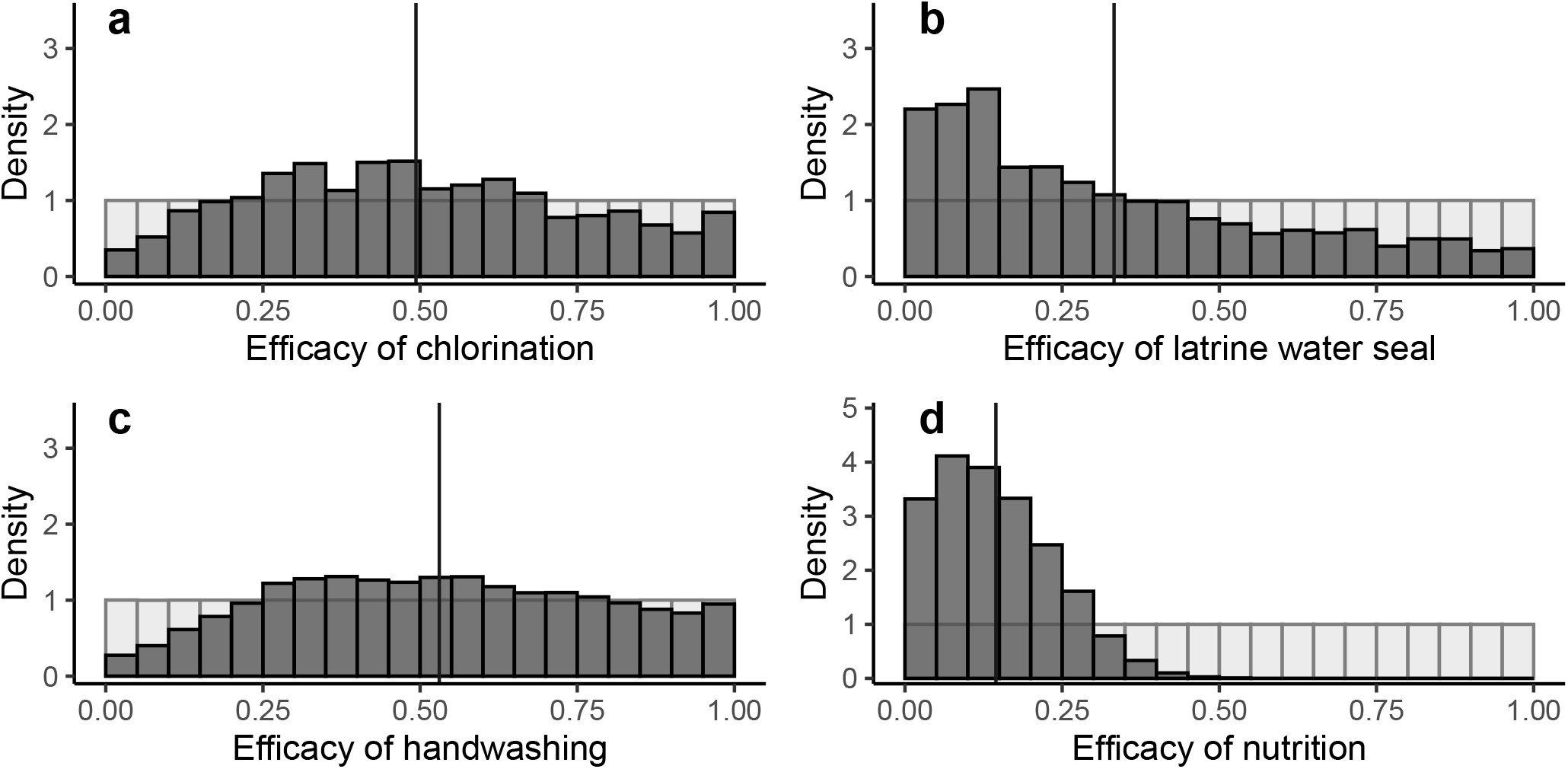
Efficacy. Posterior distributions (dark grey) of the efficacy of (a) water chlorination, (b) the fomite pathway reproduction number, (c) handwashing, and (d) the other pathway reproduction number. Prior distributions (light grey) are given for all four sampled parameters.

## Discussion

Mechanistic models are valuable hypothesis generating tools, complementing epidemiological analyses often used for hypothesis testing. They can be particularly useful alongside randomized controlled trials (RCTs), which provide rigorous assessments of a specific hypothesis but do not generalize easily to other contexts. Here, we developed a steady-state analytical framework and Bayesian parameter estimation approach that exploits both relative risk estimates and other contextual information collected in RCTs, taking advantage of the rich individual-level data in the trial. In particular, sampling-importance resampling is an ideal algorithm for developing inference from relative risk estimates or other non-epidemic disease data [24–26]. Applying this framework to a WASH RCT we found that no single pathway was likely sufficient to sustain transmission, suggesting the potential need for multiple interventions, including some not included in traditional WASH RCTs.

One important feature of our approach is the focus on steady state analysis. Although there is an extensive mathematical biology literature studying steady state properties of dynamics transmission models that focus both on stability analysis and the estimation of ℛ_0_ [27], little work has been done to build an inferential framework around steady state solutions to take advantage of epidemiological data collected in RCTs and observational studies. Instead, most inferential frameworks built around analyzing infectious disease transmission models are designed to incorporate time-series data often from passive surveillance. For instance, increasingly, studies are using transmission models with epidemic data to estimate ℛ_0_ [28, 29]. The framework that we developed here takes advantage of relative risk estimates comparing prevalence among different sub-populations that have different pathway-specific exposures [30]. RCTs provide the most rigorous data to exploit since the experimental set up eliminates confounding biases.

Pathogens that can exploit multiple environmental transmission pathways, like enteric pathogens, require a modeling framework that explicitly and distinctly incorporates these pathways [31]. Modeling multiple pathways allows us to ascertain the combination of interventions that optimally reduces diarrhea, subject to programmatic constraints. In our analysis of the WASH Benefits data, we found that there was not a single dominant environmental pathway that was likely to sustain transmission on its own, suggesting that multiple interventions would be needed to eliminate transmission. The perspective that traditional WASH interventions are not blocking all the important pathways causing infection is actively being discussed in the literature [11, 32, 33]. Our finding that no single pathway was sufficient provides empirical backing to this perspective. We also estimated that the fraction of the population covered by the study was small, likely under 10%. Although the direct effects of interventions can be estimated in RCTs with low coverage, some WASH interventions, particularly sanitation, act primarily through indirect effects. Previous work has shown that indirect effects are unlikely to be apparent until greater intervention coverage is achieved [34, 35]. Our future work will explore the potential disease reduction outcomes if the WASH Benefits interventions were implemented on a larger scale.

We also found that the reported diarrheal prevalence was consistent with a wide range of values for many of the environmental pathway strength and efficacy parameters. The practical unidentifiability of these parameters is due in part to trade-offs between the pathway strength and efficacy parameters. For example, the prevalence data may be explained by a strong water transmission pathway and weak efficacy of chlorination. Alternatively, the data may be explained by a weak water transmission pathway, regardless of the efficacy of chlorination. Additional data may be able to resolve this kind of parameter unidentifiability and would allow for more detailed insight into the multiple, complicated factors underlying diarrheal disease transmission. Particularly relevant here are the increasing number of studies collecting environmental samples from different media to inform pathways-specific exposure [36–39]. This has promise to add more specificity to exposure variables, which have traditionally been based on the presence of infrastructure (e.g., piped water or the presence of sanitation structures). Multiplex molecular technology now allows for more efficient testing of environmental samples for a wide array of pathogens. Given this new, more affordable technology, it is important to develop and standardize environmental sampling strategies for different transmission pathways in a way that complements standard epidemiological data sets. Environmental data promises to resolve many uncertainties in parameter estimation. More work is needed, both theoretical and practical, to ensure that the environmental sampling data can inform transmission pathways and, ultimately, case data. This new molecular technology can also affordably identify pathogens in stool samples, which can improve parameter estimation by replacing highly variable and uncertain natures of the self-reported diarrhea outcomes. Stool analysis also provides an enhanced understanding of infection, and not just disease, which would more directly inform our disease transmission modeling framework.

We made several assumptions in the model that could be resolved through more extensive data collection. For example, we assumed that individuals in a cluster all interact in a shared environment distinct from other environments. A more detailed characterization of the environment could allow for the relaxation of this assumption. We also assumed that the impacts of interventions could be identified from a proxy indicator and that preexisting sanitation and hygiene conditions meeting the proxy indicator were as efficacious as the intervention. Our results appear to suggest that the interventions were likely more efficacious than the preexisting WASH conditions because the model was unable to achieve the low prevalence estimates reported in those intervention arms. Future trials should consider how to measure and indicate preexisting WASH conditions as well as the fidelity of and adherence to the intervention(s). Future modeling work could also investigate separate effectiveness parameters for conditions and interventions, although that approach could substantially increase the computational burden of simulation.

## Conclusion

The strength of this work is in the integration of an advanced mathematical framework, a computational approach leading to a robust understanding of uncertainty, and the large, well-executed trial that supplied the data. Our work highlights the benefits of underutilized, interdisciplinary collaborations between mathematical epidemiologists and infectious disease trialists. This framework lays the groundwork for further analysis to better explain WASH RCT results, asking questions about completeness, i.e., the degree to which interventions block most or all transmission pathways, and how the effectiveness of the interventions may increase with increasing coverage and compliance. This framework can be used in future work to examine policy-relevant questions about completeness, coverage, and compliance, and can be adapted to a variety of other WASH and other types of RCTs. The application of mathematical modeling to estimate the impact of WASH interventions across different conditions—coverage, adherence, background infection rate, intervention efficacy—could improve external validity and deliver policy-relevant findings across contexts to better inform public infrastructure investment.

## Data Availability

The WASH Benefits Bangladesh data is publicly available at https://osf.io/tprw2/. The data and code underlying the results of this paper are available on a GitHub repository at https://github.com/afbrouwer/WASH_RCT_transmission_modeling.

https://github.com/afbrouwer/WASH_RCT_transmission_modeling

## Supporting information

**S1 Appendix. Supporting Information**. The supporting information includes the generic SISE–RCT model equations, the WASH Benefits Bangladesh SISE–RCT model equations, and supplemental results, including distributions of likelihoods, timepoint- and arm-specific relative basic reproduction numbers, and fraction of the population enrolled in the study.

## Acknowledgments

This work was funded by the Bill & Melinda Gates Foundation (www.gatesfoundation.org; grant INV-005081) and the National Science Foundation (www.nsf.gov; grant DMS-1853032). We thank the WASH Benefits investigators who supported data acquisition and initial interpretation of our work, including Benjamin Arnold, John Colford Jr, Clair Null, Amy Pickering, Andrew Mertens, Ayse Ercumen, and Jade Benjamin-Chung.

