## Supplementary material for "Leveraging infectious disease models to interpret randomized controlled trials: controlling enteric pathogen transmission through water, sanitation, and hygiene interventions": S1 Appendix

### SISE–RCT model equations and reparameterization

The full SISE–RCT model equations for a single-intervention are as follows.

$$\begin{aligned}
\frac{dS_+}{dt} &= -\left(\phi_{\beta_1}\beta_1 E_1 + \phi_{\beta_2}\beta_2 E_2 + \cdots + \phi_{\beta_n}\beta_n E_n\right) S_+ + \gamma I_+,\\
\frac{dI_+}{dt} &= \left(\phi_{\beta_1}\beta_1 E_1 + \phi_{\beta_2}\beta_2 E_2 + \cdots + \phi_{\beta_n}\beta_n E_n\right) S_+ - \gamma I_+,\\
\frac{dS_-}{dt} &= -\left(\beta_1 E_1 + \beta_2 E_2 + \cdots + \beta_n E_n\right) S_- + \gamma I_-,\\
\frac{dI_-}{dt} &= \left(\beta_1 E_1 + \beta_2 E_2 + \cdots + \beta_n E_n\right) S_- - \gamma I_-,\\
\frac{dE_1}{dt} &= \alpha_1(\phi_{\alpha_1} I_+ + I_-) - \xi_1 E_1,\\
\frac{dE_2}{dt} &= \alpha_2(\phi_{\alpha_2} I_+ + I_-) - \xi_2 E_2,\\
&\vdots\\
\frac{dE_n}{dt} &= \alpha_2(\phi_{\alpha_n} I_+ + I_-) - \xi_n E_n.
\end{aligned} \tag{S1}$$

The disease-free initial conditions of this system are $S_+ = \omega\rho + (1-\omega)\rho_0$ and $S_- = \omega(1-\rho) + (1-\omega)(1-\rho_0)$, with all other compartments 0.

Parameters $\beta$, $\alpha$, $\xi$, and $\gamma$ are not separately identifiable from steady-state data. We can see this fact by setting each $dE_i/dt$ equation in Eq. S1 to 0 (a kind of quasi-steady-state assumption), solving for the $E_i$,

$$E_i = \frac{\alpha_i}{\xi_i}(\phi_{\alpha_i} I_+ + I_-) \tag{S2}$$

and substituting those expressions into the remaining equations.

$$\begin{aligned}
\frac{dS_+}{dt} &= -\left(\phi_{\beta_1}\frac{\beta_1\alpha_1}{\xi_1}(\phi_{\alpha_1}I_+ + I_-) + \phi_{\beta_2}\frac{\beta_2\alpha_2}{\xi_2}(\phi_{\alpha_2}I_+ + I_-) + \cdots + \phi_{\beta_n}\frac{\beta_n\alpha_n}{\xi_n}(\phi_{\alpha_n}I_+ + I_-)\right) S_+ + \gamma I_+,\\
\frac{dI_+}{dt} &= \left(\phi_{\beta_1}\frac{\beta_1\alpha_1}{\xi_1}(\phi_{\alpha_1}I_+ + I_-) + \phi_{\beta_2}\frac{\beta_2\alpha_2}{\xi_2}(\phi_{\alpha_2}I_+ + I_-) + \cdots + \phi_{\beta_n}\frac{\beta_n\alpha_n}{\xi_n}(\phi_{\alpha_n}I_+ + I_-)\right) S_+ - \gamma I_+,\\
\frac{dS_-}{dt} &= -\left(\frac{\beta_1\alpha_1}{\xi_1}(\phi_{\alpha_1}I_+ + I_-) + \frac{\beta_2\alpha_2}{\xi_2}(\phi_{\alpha_2}I_+ + I_-) + \cdots + \frac{\beta_n\alpha_n}{\xi_n}(\phi_{\alpha_n}I_+ + I_-)\right) S_- + \gamma I_-,\\
\frac{dI_-}{dt} &= \left(\frac{\beta_1\alpha_1}{\xi_1}(\phi_{\alpha_1}I_+ + I_-) + \frac{\beta_2\alpha_2}{\xi_2}(\phi_{\alpha_2}I_+ + I_-) + \cdots + \frac{\beta_n\alpha_n}{\xi_n}(\phi_{\alpha_n}I_+ + I_-)\right) S_- - \gamma I_-.
\end{aligned} \tag{S3}$$

When the resulting equations are at steady-state (i.e., setting each equation to 0), for each pathway $i$, the 4 pathway-specific parameters are only found in the identifiable parameter combination $\mathcal{R}_{0,i} = \frac{\beta_i\alpha_i}{\gamma_i\xi_i}$. The system of equations Eq. (3), therefore, have the same steady state solutions as Eq. (S3).

#### WASH Benefits Bangladesh SISE–RCT model equations

The equations for the WASH Benefits Bangladesh model are as follows. For brevity, we omit the $dI/dt$ equations, each of which is given by $dI/dt = -dS/dt$. The subscripts $i$ on the population variables indicate the associated adherence group.

$$
\begin{aligned}
\frac{dS_0}{dt} &= -S_0\left(\mathcal{R}_{0,w}E_w + \mathcal{R}_{0,f}E_f + \mathcal{R}_{0,o}E_o\right) + I_0 \\
\frac{dS_{\mathrm{W}}}{dt} &= -S_{\mathrm{W}}\left(\phi_{\beta_w,\mathrm{W}}\mathcal{R}_{0,w}E_w + \mathcal{R}_{0,f}E_f + \mathcal{R}_{0,o}E_o\right) + I_{\mathrm{W}} \\
\frac{dS_{\mathrm{S}}}{dt} &= -S_{\mathrm{S}}\left(\phi_{\alpha_w,\mathrm{S}}\mathcal{R}_{0,w}E_w + \mathcal{R}_{0,f}E_f + \mathcal{R}_{0,o}E_o\right) + I_{\mathrm{S}} \\
\frac{dS_{\mathrm{H}}}{dt} &= -S_{\mathrm{H}}\left(\mathcal{R}_{0,w}E_w + \phi_{\beta_f,\mathrm{H}}\mathcal{R}_{0,f}E_f + \mathcal{R}_{0,o}E_o\right) + I_{\mathrm{H}} \\
\frac{dS_{\mathrm{N}}}{dt} &= -S_{\mathrm{N}}\phi_{\beta,\mathrm{N}}\left(\mathcal{R}_{0,w}E_w + \mathcal{R}_{0,f}E_f + \mathcal{R}_{0,o}E_o\right) + I_{\mathrm{N}} \\
\frac{dS_{\mathrm{WS}}}{dt} &= -S_{\mathrm{WS}}\left(\phi_{\beta_w,\mathrm{W}}\phi_{\alpha_w,\mathrm{S}}\mathcal{R}_{0,w}E_w + \mathcal{R}_{0,f}E_f + \mathcal{R}_{0,o}E_o\right) + I_{\mathrm{WS}} \\
\frac{dS_{\mathrm{WH}}}{dt} &= -S_{\mathrm{WH}}\left(\phi_{\beta_w,\mathrm{W}}\mathcal{R}_{0,w}E_w + \phi_{\beta_f,\mathrm{H}}\mathcal{R}_{0,f}E_f + \mathcal{R}_{0,o}E_o\right) + I_{\mathrm{WH}} \\
\frac{dS_{\mathrm{WN}}}{dt} &= -S_{\mathrm{WN}}\phi_{\beta,\mathrm{N}}\left(\phi_{\beta_w,\mathrm{W}}\mathcal{R}_{0,w}E_w + \mathcal{R}_{0,f}E_f + \mathcal{R}_{0,o}E_o\right) + I_{\mathrm{WN}} \\
\frac{dS_{\mathrm{SH}}}{dt} &= -S_{\mathrm{SH}}\left(\phi_{\alpha_w,\mathrm{S}}\mathcal{R}_{0,w}E_w + \phi_{\beta_f,\mathrm{H}}\mathcal{R}_{0,f}E_f + \mathcal{R}_{0,o}E_o\right) + I_{\mathrm{SH}} \\
\frac{dS_{\mathrm{SN}}}{dt} &= -S_{\mathrm{SN}}\phi_{\beta,\mathrm{N}}\left(\phi_{\alpha_w,\mathrm{S}}\mathcal{R}_{0,w}E_w + \mathcal{R}_{0,f}E_f + \mathcal{R}_{0,o}E_o\right) + I_{\mathrm{SN}} \\
\frac{dS_{\mathrm{HN}}}{dt} &= -S_{\mathrm{HN}}\phi_{\beta,\mathrm{N}}\left(\mathcal{R}_{0,w}E_w + \phi_{\beta_f,\mathrm{H}}\mathcal{R}_{0,f}E_f + \mathcal{R}_{0,o}E_o\right) + I_{\mathrm{HN}} \\
\frac{dS_{\mathrm{WSH}}}{dt} &= -S_{\mathrm{WSH}}\left(\phi_{\beta_w,\mathrm{W}}\phi_{\alpha_w,\mathrm{S}}\mathcal{R}_{0,w}E_w + \phi_{\beta_f,\mathrm{H}}\mathcal{R}_{0,f}E_f + \mathcal{R}_{0,o}E_o\right) + I_{\mathrm{WSH}} \\
\frac{dS_{\mathrm{WSN}}}{dt} &= -S_{\mathrm{WSN}}\left(\phi_{\beta_w,\mathrm{W}}\phi_{\alpha_w,\mathrm{S}}\mathcal{R}_{0,w}E_w + \mathcal{R}_{0,f}E_f + \mathcal{R}_{0,o}E_o\right) + I_{\mathrm{WSN}} \\
\frac{dS_{\mathrm{WHN}}}{dt} &= -S_{\mathrm{WHN}}\phi_{\beta,\mathrm{N}}\left(\phi_{\beta_w,\mathrm{W}}\mathcal{R}_{0,w}E_w + \phi_{\beta_f,\mathrm{H}}\mathcal{R}_{0,f}E_f + \mathcal{R}_{0,o}E_o\right) + I_{\mathrm{WHN}} \\
\frac{dS_{\mathrm{SHN}}}{dt} &= -S_{\mathrm{SHN}}\phi_{\beta,\mathrm{N}}\left(\phi_{\alpha_w,\mathrm{S}}\mathcal{R}_{0,w}E_w + \phi_{\beta_f,\mathrm{H}}\mathcal{R}_{0,f}E_f + \mathcal{R}_{0,o}E_o\right) + I_{\mathrm{SHN}} \\
\frac{dS_{\mathrm{WSHN}}}{dt} &= -S_{\mathrm{WSHN}}\phi_{\beta,\mathrm{N}}\left(\phi_{\beta_w,\mathrm{W}}\phi_{\alpha_w,\mathrm{S}}\mathcal{R}_{0,w}E_w + \phi_{\beta_f,\mathrm{H}}\mathcal{R}_{0,f}E_f + \mathcal{R}_{0,o}E_o\right) + I_{\mathrm{WSHN}}
\end{aligned}
\tag{S4}
$$

#### Supplemental results

The negative log-likelihood distributions of the prior (parameter samples generated to uniform on their spans) and the posterior (parameter samples resampled from the prior sample set based on the negative-log likelihood) parameter samples are given in Fig. S1. This figure demonstrates that the resampling procedure successfully generated a posterior with an improved distribution of fits to the data.

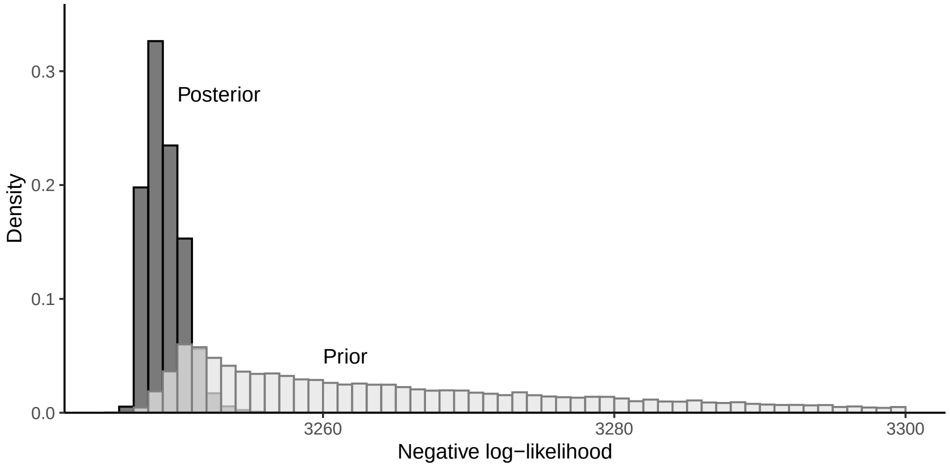


**Fig S1.** Distribution of negative log-likelihood fits to the data for parameter samples in the prior and posterior sample sets.

A plot comparing the data by wave (not combining midline and endline) to the posterior distributions of simulated prevalence is given below.

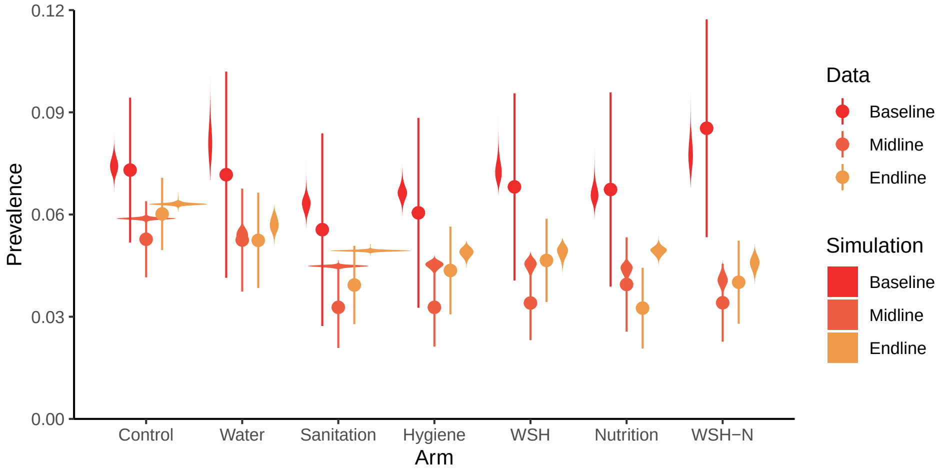


**Fig S2.** Prevalence of self-reported diarrhea (7-day recall) in WASH Benefits comparing the baseline (red) to the combined midline (orange) and endline (yellow) surveys, as well as posterior distributions of simulated prevalence (violine plots).

The estimates of relative timepoint- and arm-specific basic reproduction numbers are given in Figure S3. Both the midline and endline surveys were associated with lower reproduction numbers than the baseline survey, for all arms. The S and H arms were significantly different at $\alpha$=0.05 from the control arm across all time points. The W and WSHN arms were significantly from at $\alpha$=0.1 from the control arm across all time points. The WSH and N arms were not significantly different from the control arm.

The posterior distribution of estimated fraction of the population enrolled in the study is given in Fig. S4.

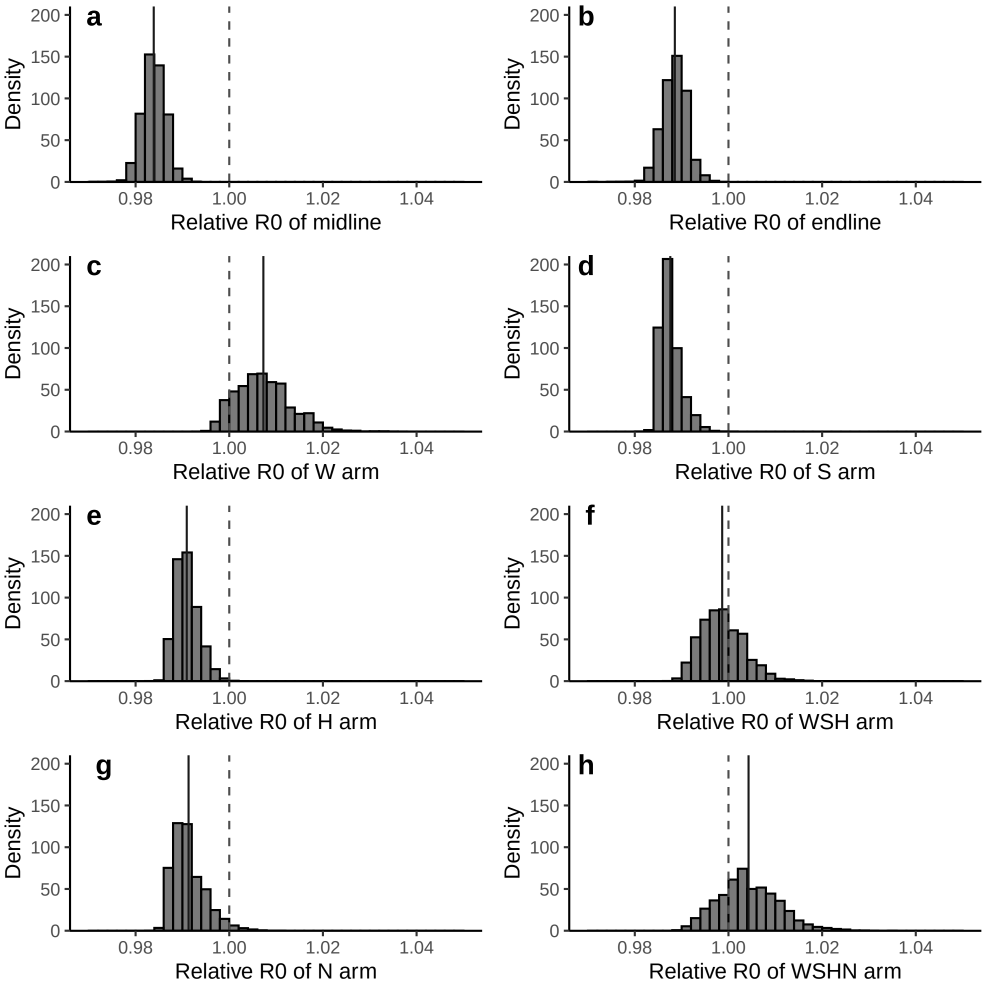


**Fig S3.** Distribution of timepoint- and arm-specific relative basic reproduction numbers ($\mathcal{R}_0$). The dotted line corresponds to 1.00, or no difference from the control arm at baseline. The solid lines give the mean values of the distributions.

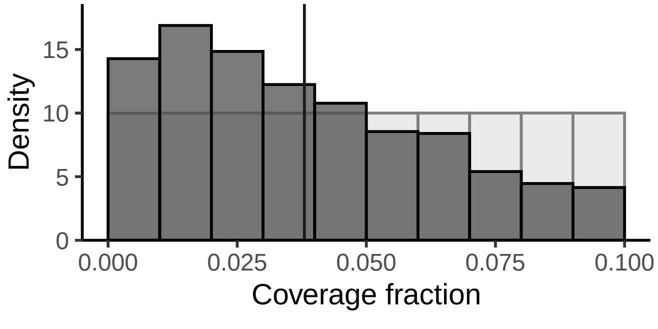


**Fig S4.** Posterior (grey) and prior (white) distributions for the estimated fraction of the population enrolled in the study.
